# Early Prediction of Hypertensive Disorders of Pregnancy Using Machine Learning and Medical Records from the First and Second Trimesters

**DOI:** 10.1101/2024.11.21.24317720

**Authors:** Seyedeh Somayyeh Mousavi, Kim Tierney, Chad Robichaux, Sheree Lynn Boulet, Cheryl Franklin, Suchitra Chandrasekaran, Reza Sameni, Gari D. Clifford, Nasim Katebi

## Abstract

Hypertensive disorders of pregnancy (HDPs) remain a major challenge in maternal health. Early prediction of HDPs is crucial for timely intervention. Most existing predictive machine learning (ML) models rely on costly methods like blood, urine, genetic tests, and ultrasound, often extracting features from data gathered throughout pregnancy, delaying intervention.

This study developed an ML model to identify HDP risk before clinical onset using affordable methods. Features were extracted from blood pressure (BP) measurements, body mass index values (BMI) recorded during the first and second trimesters, and maternal demographic information.

We employed a random forest classification model for its robustness and ability to handle complex datasets. Our dataset, gathered from large academic medical centers in Atlanta, Georgia, United States (2010-2022), comprised 1,190 patients with 1,216 records collected during the first and second trimesters. Despite the limited number of features, the model’s performance demonstrated a strong ability to accurately predict HDPs. The model achieved an F1- score, accuracy, positive predictive value, and area under the receiver-operating characteristic curve of 0.76, 0.72, 0.75, and 0.78, respectively.

In conclusion, the model was shown to be effective in capturing the relevant patterns in the feature set necessary for predicting HDPs. Moreover, it can be implemented using simple devices, such as BP monitors and weight scales, providing a practical solution for early HDPs prediction in low-resource settings with proper testing and validation. By improving the early detection of HDPs, this approach can potentially help with the management of adverse pregnancy outcomes.

## I. INTRODUCTION

Hypertensive disorders of pregnancy (HDPs) are a leading cause of maternal and fetal morbidity and mortality worldwide, posing a significant risk to maternal and infant health [1]. Elevated blood pressure (BP), protein in the urine, severe headaches, and vision changes are some of the main symptoms of these disorders [2]–[4]. In 2019, the Institute for Health Metrics and Evaluation (IHME) Global Burden of Disease (GBD) reported that the global mean prevalence of HDPs is 116.4 per 100,000 women of childbearing age [5]. In the United States, HDPs are common pregnancy complications. Between 2017 and 2019, the prevalence of HDPs among delivery hospitalizations increased from 13.3% to 15.0%. Notably, 31.6% of deaths occurring during delivery hospitalizations involved HDPs [6].

In a normotensive pregnancy, BP typically follows a specific pattern reflecting physiological changes to support the developing fetus [7], [8]. A typical pregnancy lasts around 40 weeks, with a common range from 37 to 42 weeks [9], [10]. Health care providers divide this period into three stages called trimesters, each lasting about three months, to mark distinct stages of fetal development [11], [12]. During the early first trimester, BP usually remains stable or slightly decreases compared to pre-pregnancy levels. As the pregnancy advances into the second trimester, BP generally stays within the normal range but often begins to rise by the mid-second trimester. In the third trimester, BP continues to increase gradually due to the growing demands of the fetus. During labor and delivery, BP may fluctuate but should return to normal levels after childbirth [13].

Studies by Harper et al. [14] and Macdonald-Wallis et al. [15] support this pattern of BP changes throughout pregnancy. Their datasets involved 300 and 13,016 pregnant women, respectively. According to the findings, systolic blood pressure (SBP) and diastolic blood pressure (DBP) show mean variations of less than 10 mmHg during pregnancy. The mean SBP increases from about 110 mmHg to more than 115 mmHg and the mean DBP changes from more than 65 mmHg to about 70 mmHg. Therefore, accurate analysis of BP during pregnancy is essential due to its substantial impact on the health and well-being of both the mother and the fetus. Precise

BP readings enable physicians to make informed decisions regarding the management of HDPs [16]. A notable study by Gunderson et al. [17] assessed the risk factors for HDPs in pregnant women by analyzing a comprehensive BP dataset comprising 249,892 individuals and examining BP patterns during pregnancy. They classified BP values from the first half of pregnancy into six specific groups and fitted models using a third-order polynomial curve for each category. Subsequently, they calculated the risk factors for HDPs for pregnant women based on the best fit of their early BP values in one of the models, along with demographic features.

In addition to BP patterns, various maternal factors, including Body Mass Index (BMI) and demographic features, are associated with HDPs. Understanding these factors alongside BP patterns is essential for the early detection of HDPs, which is crucial for improving maternal and fetal health outcomes. In the following sections, we will examine the relationship between these factors and HDPs.

### A. Hypertensive Disorders of Pregnancy

The International Classification of Diseases, Ninth Revision, Clinical Modification (ICD-9-CM), and the newer version, Tenth Revision, Clinical Modification (ICD-10-CM) are standardized coding systems for classifying diagnoses, symptoms, and medical procedures in healthcare. These systems include specific codes and descriptions for various hypertension groups, as shown in Tables I and II. The main difference between these codes is the level of details they provide for diagnoses and procedures. Within the ICD-9-CM classification, HDPs are categorized into four groups [4]: Gestational hypertension (GE), Preeclampsia/Eclampsia, Pre-existing (chronic) hypertension, and Unspecified hypertension.

**TABLE I:** ICD-9-CM Diagnosis Codes for Hypertension in Pregnancy [4].

| Type of Hypertension | Description | Code |
| --- | --- | --- |
| **Preexisting** | Benign essential hypertension complicating pregnancy, childbirth, and the puerperium | 642.0x |
|  | Hypertension secondary to renal disease, complicating pregnancy, childbirth, and the puerperium | 642.1x |
|  | Other pre-existing hypertension complicating pregnancy, childbirth, and the puerperium | 642.2x |
| **Gestational** | Transient hypertension of pregnancy | 642.3x |
| **Preeclampsia/eclampsia** | Mild or unspecified pre-eclampsia | 642.4x |
|  | Severe pre-eclampsia | 642.5x |
|  | Eclampsia | 642.6x |
|  | Pre-eclampsia or eclampsia superimposed on pre-existing hypertension | 642.7x |
| **Unspecified** | Unspecified hypertension complicating pregnancy, childbirth, and the puerperium | 642.9x |

**TABLE II:** General structure of ICD-10-CM Diagnosis Codes for Hypertension in Pregnancy [18].

| Type of Hypertension | Description | Code |
| --- | --- | --- |
| **Preexisting** | Preexisting hypertension complicating pregnancy, childbirth and the puerperium | 010.x |
| **Preexisting and Preeclampsia** | Pre-existing hypertension with pre-eclampsia | 011.x |
| **Gestational** | Gestational [pregnancy-induced] edema and proteinuria without hypertension | 012.x |
|  | Gestational [pregnancy-induced] hypertension without significant proteinuria | 013.x |
| **Preeclampsia/eclampsia** | Pre-eclampsia | 014.x |
|  | Eclampsia | 015.x |
| **Unspecified** | Unspecified maternal hypertension | 016.x |

**TABLE III:** Total gestational weight gain and weekly rate of weight gain recommendations regarding pre-pregnancy BMI groups [38].

| Pre-Pregnancy Weight Group | BMI Ranges (kg/m<sup>2</sup>) | Total Weight Gain (kg) | Weekly Rate of Weight Gain (kg) |
| --- | --- | --- | --- |
| Underweight | ≤ 18.5 | 12.5 - 18 | 0.51 (0.44 - 0.58) |
| Normal weight | 18.5 - 24.9 | 11.5 - 16 | 0.42 (0.35 - 0.50) |
| Overweight | 25.0 - 29.9 | 7 - 11.5 | 0.28 (0.23 - 0.33) |
| Obese | ≥ 30.0 | 5 - 9 | 0.22 (0.17 - 0.27) |

**TABLE IV:** Distribution of Patients and Records in All Trimesters Based on the Hypertension in Pregnancy Diagnosis Labels.

| Trimester | N_Patients | N_Records | Normal | Preeclampsia/Eclampsia | Gestational | Preexisting | Unspecified |
| --- | --- | --- | --- | --- | --- | --- | --- |
| First | 2439 | 2579 | 1016 (39.40%) | 374 (14.50%) | 331 (12.83%) | 607 (23.54%) | 251 (9.73%) |
| Second | 2420 | 2555 | 646 (25.28%) | 472 (18.47%) | 409 (16.01%) | 707 (27.67%) | 321 (12.56%) |
| Third | 2183 | 2297 | 443 (19.29%) | 470 (20.46%) | 422 (18.37%) | 642 (27.95%) | 320 (13.93%) |
| Both first and second | 1927 | 2021 | 534 (26.42%) | 358 (17.71%) | 324 (16.03%) | 566 (28.01%) | 239 (11.83%) |

**TABLE V:** Distribution of Patients and Records in the First Two Trimesters Based on Normal and HDPs Diagnosis Labels.

| Trimester | N_Patients | N_Records | Normal | HDPs |
| --- | --- | --- | --- | --- |
| First | 1667 | 1721 | 1016 (59.04%) | 705 (40.96%) |
| Second | 1492 | 1527 | 646 (42.31%) | 881 (57.69%) |
| Both first and second | 1190 | 1216 | 534 (43.91%) | 682 (56.09%) |

**TABLE VI:** Distribution of Records Based on the Age and Diagnosis Labels.

| Characteristics | N.All | N.HDPs | N.Normal |
| --- | --- | --- | --- |
| Maternal age, y, mean (SD) | 32.01±5.36 | 31.7±5.42 | 32.4±5.26 |
| **Age categories** |  |  |  |
| 16–24 | 146 (12.0%) | 94 (13.8%) | 52 (9.7%) |
| 25–29 | 304 (25.0%) | 167 (24.5%) | 137 (25.7%) |
| 30–34 | 444 (36.5%) | 262 (38.4%) | 182 (34.1%) |
| 35–39 | 268 (22.1%) | 136 (19.9%) | 132 (24.8%) |
| 40–56 | 53 (4.4%) | 22 (3.2%) | 31 (5.8%) |

**TABLE VII:** BMI distribution of records during the first and second trimesters.

| BMI Category | Trimester 1 |  |  | Trimester 2 |  |  |
| --- | --- | --- | --- | --- | --- | --- |
|  | N.All = 1216 | N.HDPs = 682 | N.Normal = 534 | N.All = 1216 | N.HDPs = 682 | N.Normal = 534 |
| Missing values | 62 (5.1%) | 32 (4.7%) | 30 (5.6%) | 45 (3.7%) | 16 (2.3%) | 29 (5.4%) |
| Underweight (< 18.5) | 14 (1.2%) | 7 (1.0%) | 7 (1.3%) | 7 (0.6%) | 2 (0.3%) | 5 (0.9%) |
| Normal weight (18.5–24.9) | 440 (36.2%) | 222 (32.6%) | 218 (40.8%) | 368 (30.3%) | 172 (25.2%) | 196 (36.7%) |
| Overweight (25–29.9) | 321 (26.4%) | 187 (27.4%) | 134 (25.1%) | 354 (29.1%) | 198 (29.0%) | 156 (29.2%) |
| Obesity class I (30–34.9) | 189 (15.5%) | 110 (16.1%) | 79 (14.8%) | 209 (17.2%) | 135 (19.8%) | 74 (13.9%) |
| Obesity class II (35–39.9) | 108 (8.9%) | 71 (10.4%) | 37 (6.9%) | 136 (11.2%) | 92 (13.5%) | 44 (8.2%) |
| Obesity class III ($\geq 40$) | 82 (6.7%) | 53 (7.8%) | 29 (5.4%) | 97 (8.0%) | 67 (9.8%) | 30 (5.6%) |

**TABLE VIII:** Parameters of the Machine Learning Models.

| Models | Parameters |
| --- | --- |
| KNN | n_neighbors=5, weights="uniform", algorithm="auto", leaf_size=30, Power=2, metric="minkowski" |
| SVM | C=1.0, kernel='rbf', degree=3, gamma='scale', coef0=0.0, shrinking=True, probability=False, tol=0.001, cache_size=200, verbose=False, max_iter=-1, decision_function_shape='ovr' |
| MLP | hidden_layer_sizes=(100,), activation='relu', solver='adam', alpha=0.0001, batch_size='auto', learning_rate='constant', learning_rate_init=0.001, power_t=0.5, max_iter=200, shuffle=True, tol=0.0001, momentum=0.9, nesterovs_momentum=True, validation_fraction=0.1, beta_1=0.9, beta_2=0.999, epsilon=1e-08, n_iter_no_change=10, max_fun=15000 |
| XGBoost | max_depth=3, learning_rate=0.1, n_estimators=100, silent=True, objective='binary:logistic', booster='gbtree', n_jobs=1, gamma=0, min_child_weight=1, max_delta_step=0, subsample=1, colsample_bytree=1, colsample_bylevel=1, reg_alpha=0, reg_lambda=1, scale_pos_weight=1, base_score=0.5 |
| RF | n_estimators=100, criterion='gini', min_samples_split=2, min_samples_leaf=1, max_features='sqrt', bootstrap=True, verbose=0, ccp_alpha=0.0 |

**TABLE XI:**
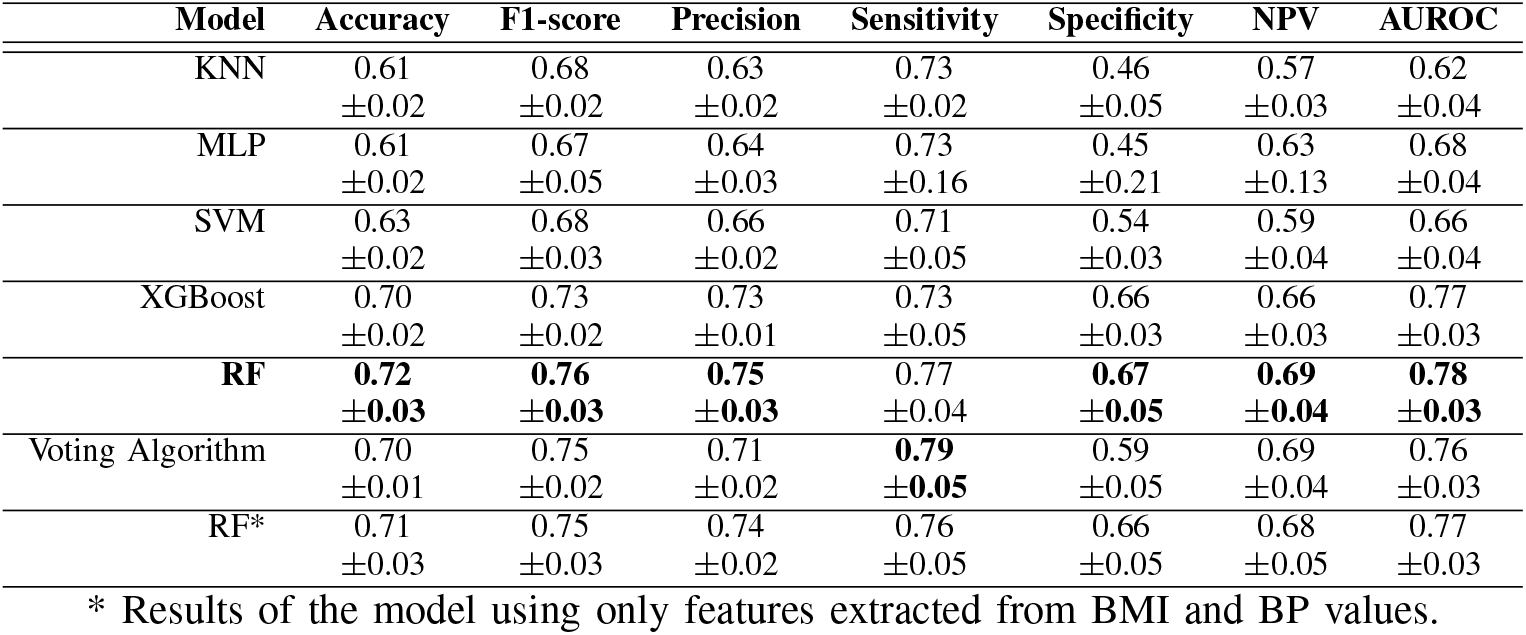
Comparison of various machine learning models for predicting HDPs versus normal pregnancies in this project.

**TABLE X:** Comparison of the best classifier for predicting HDPs in this study with those from other studies rely on costly methods, such as blood and urine tests, ultrasound, and genetic tests, and often extract features from data gathered throughout the entire gestational term of pregnancy, causing delays in the optimal time for medical intervention.

| Studies | Ges-Age | N.F | F1-score | Accuracy | Precision | Sensitivity | Specificity | NPV | AUROC |
| --- | --- | --- | --- | --- | --- | --- | --- | --- | --- |
| The best model of this research | (1,28) | 18 | 0.76<br>$\pm 0.03$ | 0.72<br>$\pm 0.03$ | 0.75<br>$\pm 0.03$ | 0.77<br>$\pm 0.04$ | 0.67<br>$\pm 0.05$ | 0.69<br>$\pm 0.04$ | 0.78<br>$\pm 0.03$ |
| Yi-xin Li et al. (2021) [42] | 2nd trimester | 38 | 0.57 | 0.92 | 0.45 | 0.79 | - | - | 0.96 |
| Leon J. Schmidt et al. (2022) [41] | (23-35) | 114 | 0.76 | 0.89 | - | 0.66 | 0.97 | 0.89 | 0.82 |
| Shilong Li et al. (2022) [56] | (1-37) | 78 | - | - | 0.16 | - | 0.95 | - | 0.87 |
| Muhlis Tahir et al. (2018) [55] | (1-37) | 17 | 0.90 | 0.95 | 0.90 | 0.90 | - | - | - |
| Yequn Chen et al. (2023) [45] | (1-14) | 11 | - | - | - | 0.65 | 0.85 | - | 0.80 |

The primary distinction between GE and preeclampsia/eclampsia, and pre-existing hypertension is the timing of onset [3]. Hypertension identified before 20 weeks of gestational age (GA) is considered pre-existing. If hypertension is detected after 20 weeks’ gestation, it is classified as either GE or preeclampsia/eclampsia. Hypertension is defined as the SBP *≥* 140 mmHg or DBP *≥* 90 mmHg. Each type of hypertension has specific characteristics and implications for maternal and fetal health. In the following, we briefly describe them [3], [4]:

- **Gestational hypertension (GE)** is defined as hypertension occurring after 20 weeks of GA without proteinuria or end-organ damage.
- **Preeclampsia (PE)** is likely in the pathway and progression to GE but it is often associated with high levels of protein in the urine and includes signs of end-organ damage or uteroplacental dysfunction.
- **Eclampsia** is a severe form of preeclampsia characterized by seizures.
- **Pre-existing (chronic) hypertension** is defined as persistent hypertension present before pregnancy or diagnosed before 20 weeks of GA.
- **Unspecified hypertension** refers to patients with insufficient information to classify hypertension into one of the specified categories.

In the ICD-10-CM code classification, there is one additional group compared to ICD-9-CM: “Pre-existing and Preeclampsia”. This new category includes patients with preeclampsia or eclampsia superimposed on pre-existing hypertension. The additional detail provided by ICD-10-CM codes allows for straightforward conversion to ICD-9-CM codes. In the next section, we describe the dataset used for this project. Due to limited access to recent versions of ICD-10-CM codes for part of the data, groups are classified based on ICD-9-CM codes, as the data was gathered before the implementation of ICD-10-CM in health centers. In cases where ICD-9-CM codes are unavailable, we use the converted version of ICD-10-CM codes to ICD-9-CM ones.

### B. Association between Demographic Features, Body Mass Index, and Hypertensive Disorders of Pregnancy

Demographic features, including sex, age, race, and ethnicity have a significant impact on BP levels and the risk of hypertension. In a recent study, we examined approximately 75 million BP values from the general population. The pre- processed BP dataset includes 1,699,955 unique individuals, consisting of 58.9% females and 41.1% males, aged 0-120 years, and spanning seven racial and ethnic groups from Georgia, USA. Our findings showed that the African American or Black group has slightly higher BP levels on average compared to the Asian, Caucasian or White, Multiple, Native Hawaiian or Other Pacific Islander, American Indian or Alaskan Native, and Hispanic racial groups [19]. The study also showed that average DBP peaks in the forties age group, whereas average SBP shows a consistent increase with age. This variation in BP across different ages is particularly relevant when considering maternal age as a determinant in the development of HDPs [20]. Specifically, women of early reproductive age and those of advanced maternal age are at higher risk for conditions such as GE and preeclampsia. The first group may lack physiological maturity leading to increased susceptibility to GE and preeclampsia [21], [22] and in the second group, there is a decline in vascular and endothelial function, which increases the risk of chronic hypertension and developing preeclampsia [23], [24]. In 2014, Ye et al. [25] conducted a study on 112,386 pregnant women in China and reported that women aged 25–29 years exhibited the lowest prevalence of HDPs, which was 4.33%. In contrast, the prevalence was significantly higher at 22.15% among those aged 35 years and older.

Maternal race and ethnicity are additional factors that may influence the incidence of HDPs [26], [27]. Previous studies have shown the impact of these factors on the development of HDPs and have incorporated them into predictive models distinguishing HDPs from normal pregnancies. In 2022, the Centers for Disease Control and Prevention (CDC) reported approximately one in five delivery hospitalizations among Black women involving HDPs [28]. A growing body of research emphasizes that structural racism, in addition to race or ethnicity, is a significant driver of disparities in the risk and outcomes of HDPs. Socioeconomic status and access to health- care further compound these inequities [29], [30]. Global data also reveal notable disparities in the HDPs burden between high-income countries and low- and middle-income countries [1]. In 2019, the highest HDP prevalence was recorded in Africa, at 334.9 per 100,000 women, followed by Southeast Asia and the Middle East, with rates of 136.8 and 121.4 per 100,000, respectively. In contrast, the Western Pacific region had the lowest prevalence, at 16.4 per 100,000 women [5], [31]. These regional statistics underscore the significant geographic disparities in HDPs.

Finally, pre-pregnancy BMI, calculated as weight in kilograms divided by the square of height in meters, and Gestational Weight Gain (GWG) are two critical factors that can affect the development of HDPs [32]. Numerous studies have reported a direct relationship between pre-pregnancy BMI and the risk of HDPs [33]–[35]. In other words, as pre-pregnancy BMI increases, the risk of incidences of HDPs rises. On the other hand, GWG is necessary to support the growing fetus and prepare the maternal body for delivery and the process of breastfeeding after that [36]. However, excessive GWG is associated with elevated BP and HDPs. Weight gain is not significant during the first trimester and usually accelerates in the second and third trimesters [37]. In 2009, the Institute of Medicine and the National Research Council published total GWG and weekly rate of weight gain recommendations during the second and third trimesters regarding pre-pregnancy BMI groups [38]. The report shows that the recommended minimum and maximum total weight gains are related to the obese and underweight BMI groups, respectively, suggesting an inverse relationship between pre-pregnancy BMI and recommended total weight gain(Table III).

### C. Aims of This Research

This study aims to classify records into two categories: Normal and HDPs, which include GE and Preeclampsia/eclampsia. Our goal is to predict HDPs in patients who initially appear to be in low-risk groups with normal blood pressure before pregnancy but later develop HDP during pregnancy. While recent advancements in machine learning (ML) and deep learning (DL) methods have led to significant progress in predicting HDPs [39], [40], the task remains an ongoing challenge. Most existing models have certain limitations that our research aims to address. The key contributions and novelties of our developed model include:

### 1) Early Prediction Focus

Our approach emphasizes early prediction of HDP during the first and second trimesters, unlike models that rely on data from the entire gestational term or the third trimester [41]–[43]. HDP symptoms typically become more apparent closer to delivery, which can enhance model performance but limits clinical usefulness for early intervention. Early detection of HDPs is vital, as it allows for timely interventions such as low-dose aspirin (LDA) therapy, one of the most effective preventive measures for HDPs. The American College of Obstetricians and Gynecologists (ACOG) recommends starting a daily dosage of 81 mg of aspirin for women at risk of HDPs between 12 and 28 weeks of gestation [44]. By early prediction of HDPs, our model can aid in optimizing management strategies and potentially reducing the risk of adverse pregnancy outcomes.

### 2) Integration of Multiple Sequential Data Points

Our model improves on previous methods by incorporating sequential measurements of SBP, DBP, and BMI from both the first and second trimesters, rather than relying on single BP or BMI values[42], [45]. This approach accounts for variations in these metrics, offering a more accurate and comprehensive understanding of maternal health during pregnancy. A single BP and BMI measurement can often be insufficient due to the numerous factors that can affect the accuracy of measurements [46]. Temporal data, which is often overlooked in existing models, is crucial for capturing the dynamic nature of BP and BMI during pregnancy, ultimately leading to more reliable predictions.

### 3) Accessible and Cost-Effectiveness

Our model is designed to be practical and accessible, especially in low-resource settings where advanced diagnostic tests may be unavailable. Many predictive models rely on features derived from costly and less accessible methods, such as blood tests, genetic tests, and Doppler ultrasound techniques [47], [47]–[52]. A literature review of 40 predictive models for GE and preeclampsia identified the four most frequently used features as BMI, uterine artery pulsatility index (UtA PI), pregnancy-associated plasma protein-A (PAPP-A), and placental growth factor (PlGF) [53]. In contrast, our approach leverages widely available and less costly metrics, including BP and BMI values and demographic features ensuring broader feasibility without compromising prediction accuracy.

In this research, we demonstrate how these contributions address the primary limitations of previous studies and enhance the predictive power of models for HDPs.

## II. METHODS

This section will discuss the dataset, preprocessing steps, feature extraction, and the ML model employed for predicting HDPs. Fig. 1 shows the graphical overview of the developed model (The study was approved by the Emory University Institutional Review Board (IRB) under protocol number STUDY00005252, with approval granted on February 2, 2024.).

**Fig 1:**
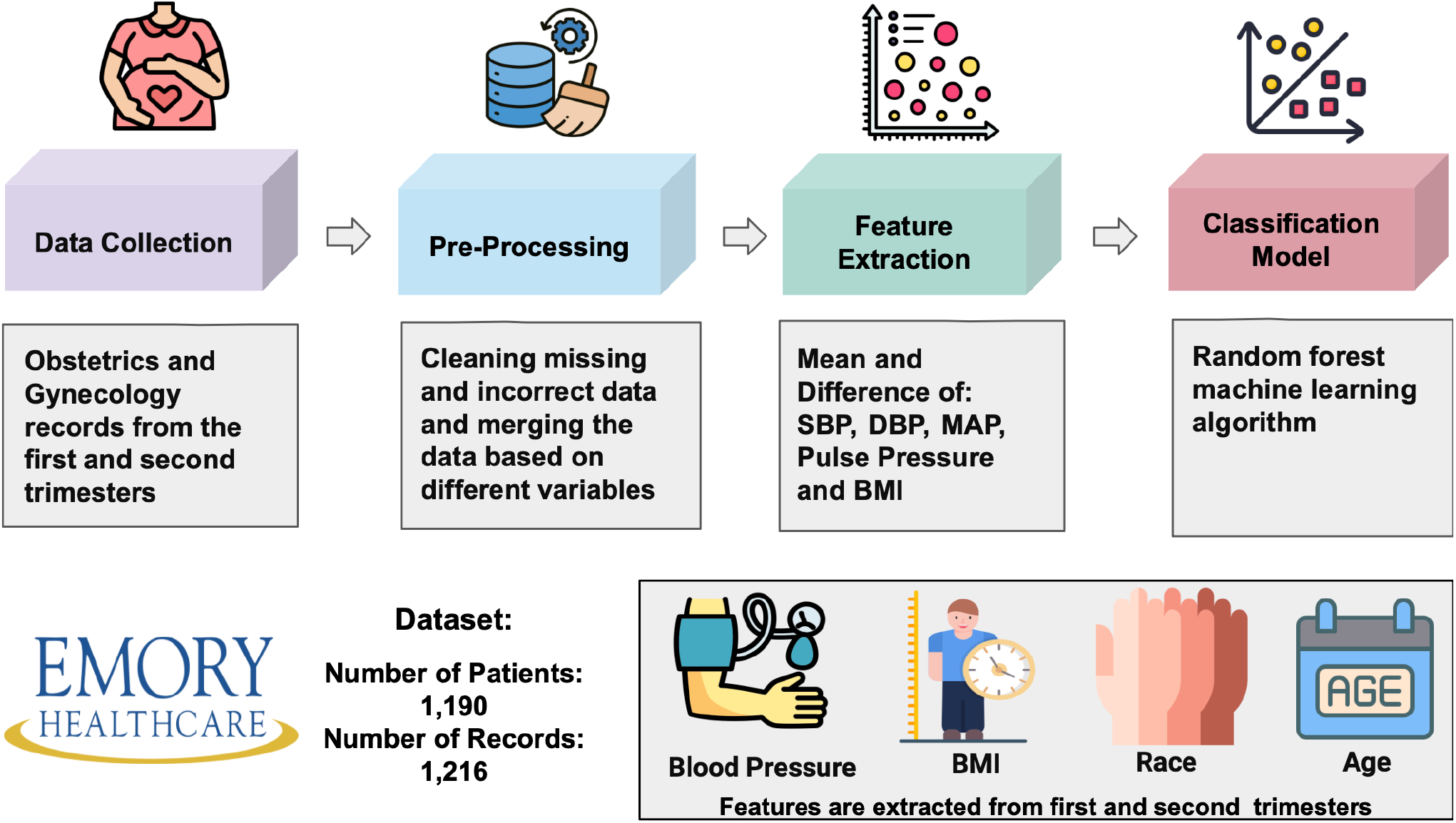
Overview of the developed model for prediction of hypertensive disorders of pregnancy using data collected from large academic medical centers in Atlanta, Georgia, United States (2010-2022). The pipeline includes data collection, preprocessing, feature extraction, and the classification model. The features consist of blood pressure and body mass index values, race, and age.

### A. Dataset

In this study, we utilized Obstetrics and Gynecology (OBGYN) medical health records collected from 2010 to 2022, over twelve years, from large academic medical centers based in Atlanta, Georgia, United States. Multiple databases were queried to form a dataset encompassing information on the Last Menstrual Period (LMP), service date, SBP, DBP, and BMI values, as well as maternal demographic features including self-reported race, and ethnicity, age, and diagnosis labels. In this project, LMP information is used to identify GA at each visit.

For labeling the records, we used both ICD-9-CM and ICD- 10-CM codes. This was necessary because the dataset was collected over an extended period, and ICD-10-CM codes were not available for a group of records. We used both coding systems to maximize the number of records included. It’s important to note that the ICD-10-CM codes can be converted to ICD-9-CM codes because it provides much information. We compared both ICD systems to ensure consistency, and in cases of discrepancies or when one code was not recorded or labeled as “unspecified hypertension” we checked the corresponding ICD-10-CM code and updated the record’s label according to the reported classification and the specific type of hypertension.

In the HDPs group, records labeled as “Pre-existing (chronic) hypertension” have hypertension before their pregnancies and are at higher risk of pregnancy complications. These women often require medical supervision throughout all trimesters. Additionally, there is a lack of information about “Unspecified hypertension”, which complicates classifying hypertension into one of the specified HDPs categories. Therefore, our approach focuses on addressing both “gestational hypertension” and “preeclampsia/eclampsia” in the HDPs group, while excluding the records with labels of “preexisting (chronic) hypertension” and “unspecified hypertension”.

### B. Data Preprocessing

The dataset contains information on 19,753 unique patients, including patient ID, service date, LMP, SBP and DBP values, birth date, and race and ethnicity. Data preprocessing involved omitting records lacking LMP information, as it is necessary to calculate GA and match diagnosis labels to a unique patient ID in the next steps. We also excluded invalid records where the service date was earlier than the LMP, resulting in a dataset of 13,139 patients. There were no missing values in date of birth recordings, but records with unknown race and ethnicity led to a final dataset of 11,393 unique patients. To exclude invalid BP values, we first removed recordings where the DBP was higher than the SBP. After addressing this issue, statistical analysis revealed that more than 99% of the BP data were within the range of DBP *≥* 40 and SBP *≥* 70. Therefore, we considered these as the BP thresholds, consistent with the preprocessing approach used in the study by P. Gunderson et al. [26]. In the next stage, we integrated the preprocessed data with the diagnosis data which includes patient ID, diagnosis date, and labels assigned by physicians based on patients’ health conditions. It is evident that as patients approach their delivery date, the diagnoses become more precise. trimester. To ensure accuracy, we selected the diagnosis label with the maximum time difference from the LMP as the final diagnosis. The time difference between the diagnosis date and LMP ranges from 1 to 294 days. After matching the relevant diagnosis labels with each record, we noticed a lack of diagnosis labels for many patients, resulting in a dataset that includes 3,077 unique patients. Then, we excluded patients who had pre-pregnancy diabetes from the dataset because diabetes is a high-risk factor during pregnancy, and patients with this condition should be under control throughout the entire pregnancy. In the final stage, we combined the preprocessed data with BMI values based on patient ID and service date. Consequently, our final dataset includes 28,664 records related to 3,025 unique patients. It should be noted that some records had missing BMI values (62 and 45 missing BMI values in the first and second trimesters, respectively). Fig. 2 shows the data preprocessing steps involved in preparing the dataset.

**Fig 2:**
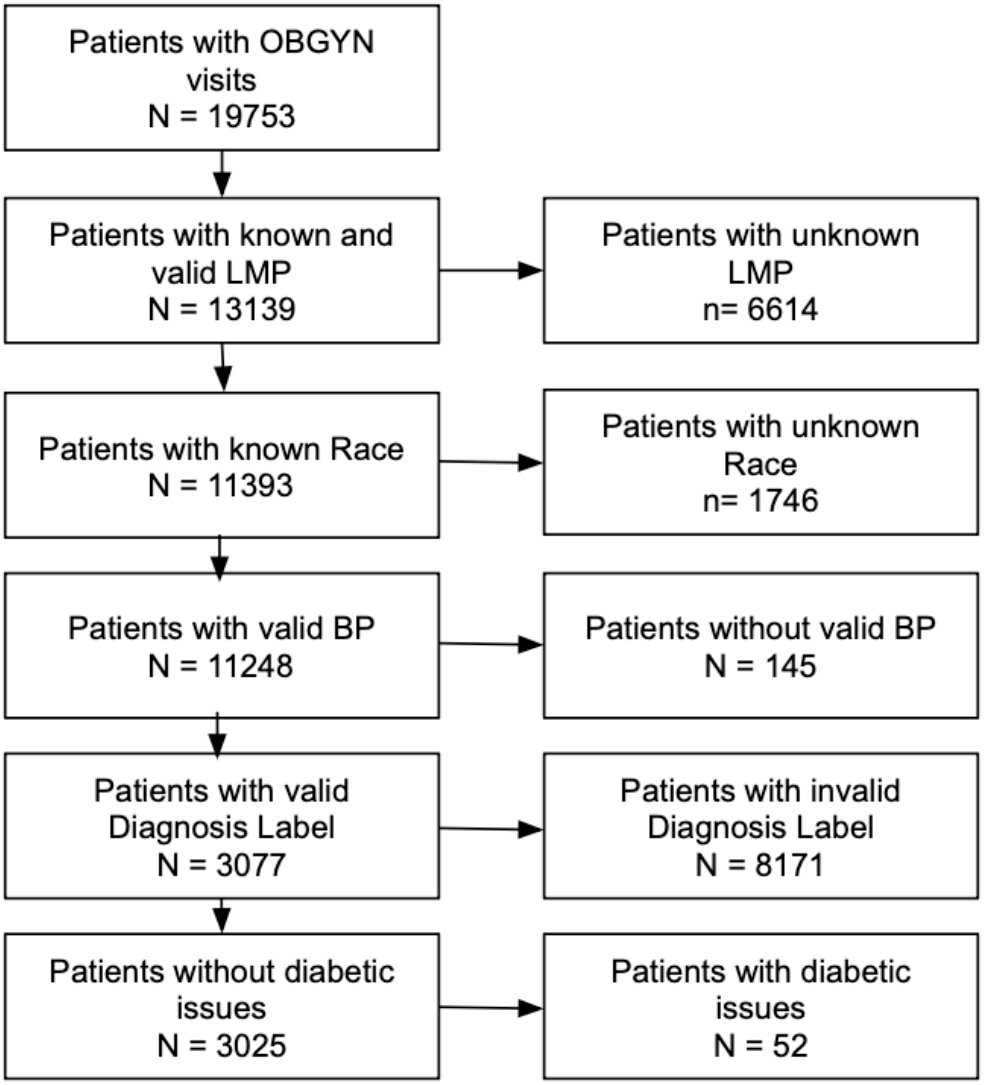
Data cleaning and preprocessing workflow. Abbreviations, OBGYIN: Obstetrics and Gynecology, LMP: Last Menstrual Period

**Fig 3:**
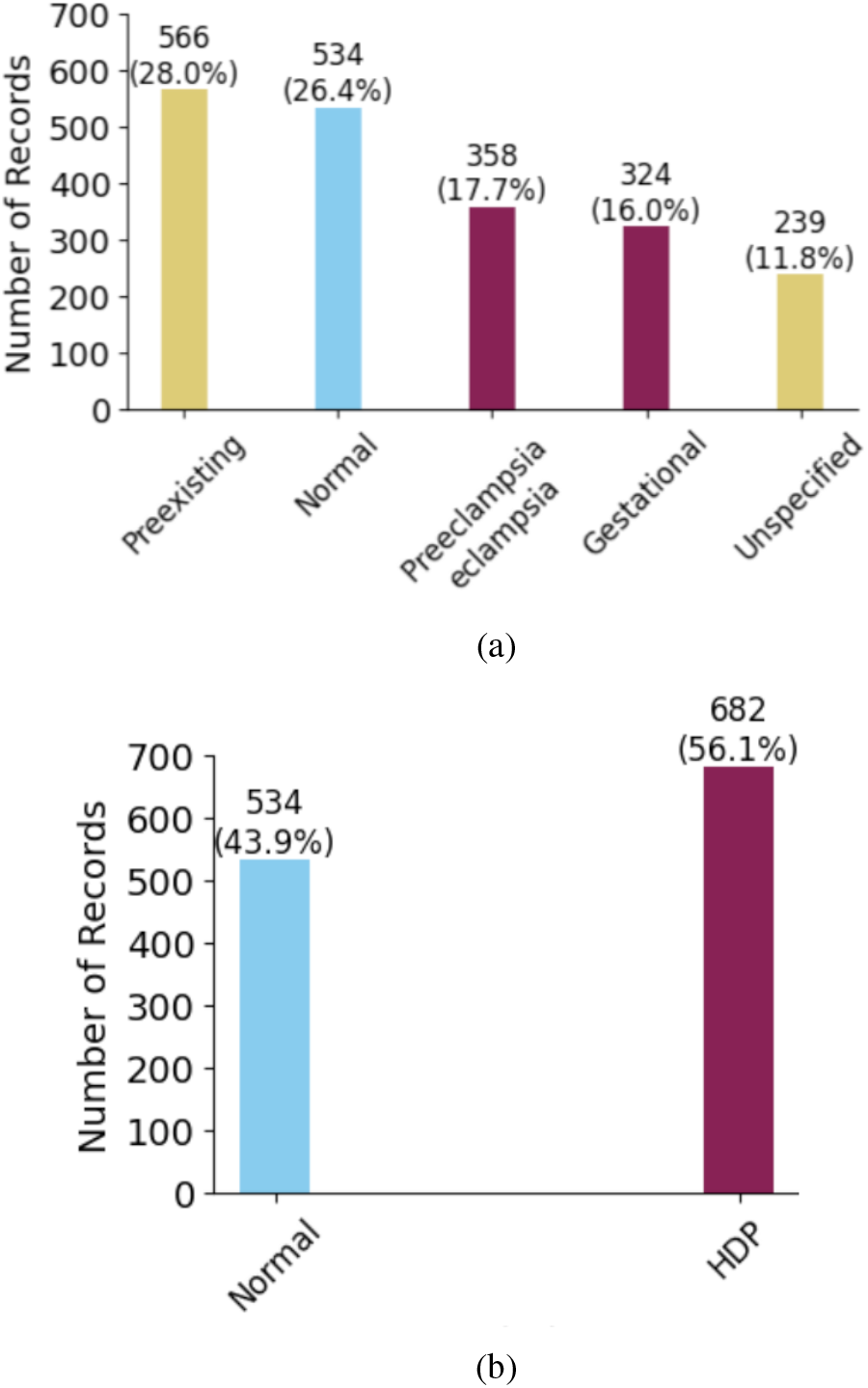
Distribution of diagnosis labels. a) Distribution of data based on Normal and hypertensive disorders of pregnancy (HDPs) diagnosis labels. b) Based on the aim of this project, records labeled as “Pre-existing (chronic) hypertension” and “Unspecified hypertension” were excluded, and the labels “Gestational Hypertension” and “Preeclampsia/Eclampsia” were renamed as HDP labels.

### C. Feature Extraction

After preprocessing the data, we calculated the GA in weeks and days by determining the difference between the LMP and the service date. GA was then categorized into trimesters as follows [54]: the first trimester was defined as 12 weeks or fewer, the second trimester as between 13 and 28 weeks, and the third trimester as 29 weeks or more. Table IV shows the distribution of patients and records across all trimesters based on the diagnosis labels. In this dataset, some records include multiple measurements within a single week, while others have missing values across several weeks. Consequently, we grouped the data by patient ID, LMP, and trimesters, and calculated mean available information of BMI and BP values across maternal demographic features including age and race. Additionally, a patient’s age during pregnancy may vary by up to one year, so we considered the minimum age during the pregnancy for each patient. Table V shows the distribution of patients and records in the first, second, and both trimesters based on the HDPs and Normal labels.

We categorized all the features considered in this study for developing the classifier for HDP vs. Normal into two groups based on their temporal characteristics: non-time-varying features, including age, race, and ethnicity, and time-varying features related to BP and BMI. In the first stage of feature extraction, in addition to SBP and DBP values, we calculated the mean arterial pressure (MAP) and pulse pressure (PP) for both the first and second trimesters. In the next step, we calculated the ratio and difference of BMI between the two trimesters as additional features. There are no missing values for any parameters except for BMI values. In total, our database contains 18 features, including mean DBP, SBP, MAP, pulse pressure, BMI values from the first and second trimesters, the differences in these values between the two trimesters, the BMI ratio (second to first trimester), as well as age and race.

### D. Feature Distributions

In this section, we examine the distribution of features in the dataset to ensure that they are appropriately represented and that any potential biases related to these features are identified.

- Non-time-varying features: Race and ethnicity are represented by six categories: Caucasian (or White), African American (or Black), Asian, American Indian (or Alaskan Native), Native Hawaiian (or Other Pacific Islander), and Hispanic. Due to the small representation of the latter three groups, which together comprise about 2 percent of the data, they were merged into a new category labeled “Multiple.” Fig. 4 illustrates the distribution of records across racial and ethnic groups, categorized by diagnosis labels. The data reveals a balanced representation within both groups. In our dataset, maternal age ranges from 16 to 56 years. We categorized ages into 5-year intervals and merged groups with less than 10 percent representation. Table VI shows the statistical distribution of age groups across both diagnosis labels. The age group 30-34 years represents the largest proportion in both Normal and HDPs groups, while the 40-56 years group represents the smallest proportion. Overall, the distribution appears balanced between the two diagnosis labels, suggesting no significant bias or trend toward any particular age group.
- Time-varying features: Features extracted from BP measurements include mean SBP, mean DBP, mean MAP, and mean PP in each trimester as well as the difference between the two trimesters. Fig. 5 demonstrates the 95% percentile range of BP distributions for the HDPs and Normal groups during the first and second trimesters. In the Normal group, SBP and DBP values tend to decrease slightly in the second trimester compared to the first, consistent with known trends in BP fluctuation throughout pregnancy. In contrast, the HDPs group shows less decrease in BP, indicating a difference in BP changes between the two groups.

**Fig 4:**
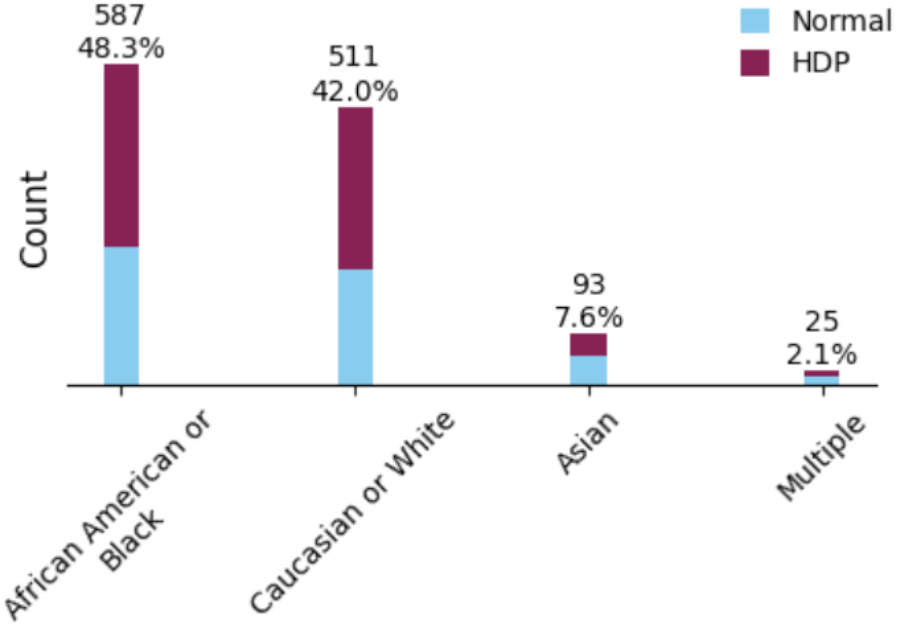
Distribution of race and ethnic groups based on diagnosis labels

**Fig 5:**
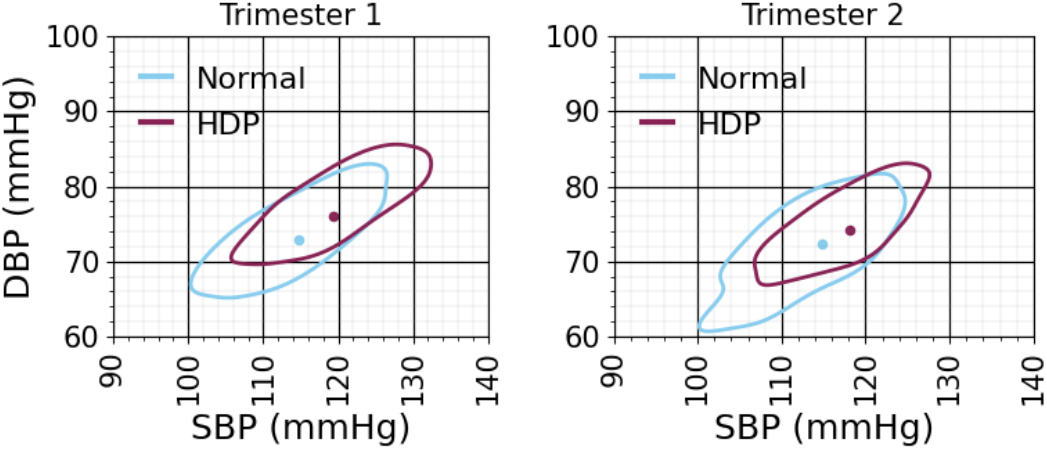
Blood pressure distributions based on the diagnosis labels. The contours correspond to the 95% percentile range of the data. Dots show the mean SBP and DBP corresponding to the diagnostic labels.

The extracted features from BMI include mean BMI during the first and second trimesters, as well as the ratio and difference. BMI is the variable in the dataset with missing values, with over 99% of records falling within the 16.0 to 56.0 range across trimesters. Additionally, more than 99% and 98% of records fall within the BMI ratio range of 0.65 to 1.55 and the BMI difference range of -6 to 6, receptively. We used these as thresholds, marking outliers as missing. The observed ratios ranged from 0.86 to 1.27, differences from -4.9 to 5.5, and BMI values from 17.1 to 56.0. Table VII shows BMI group distribution across both trimesters, with most patients in the Normal and Overweight categories. The data show that individuals with normal outcomes may belong to higher BMI groups, while those with HDPs can also be in lower BMI categories. This balance underscores the importance of retaining records with missing BMI, as weight measurements are sometimes missed in pregnancy visits. Fig. 6 further illustrates that patients with HDPs tend to have BMI ratios above one, while those with normal outcomes have ratios closer to one.

**Fig 6:**
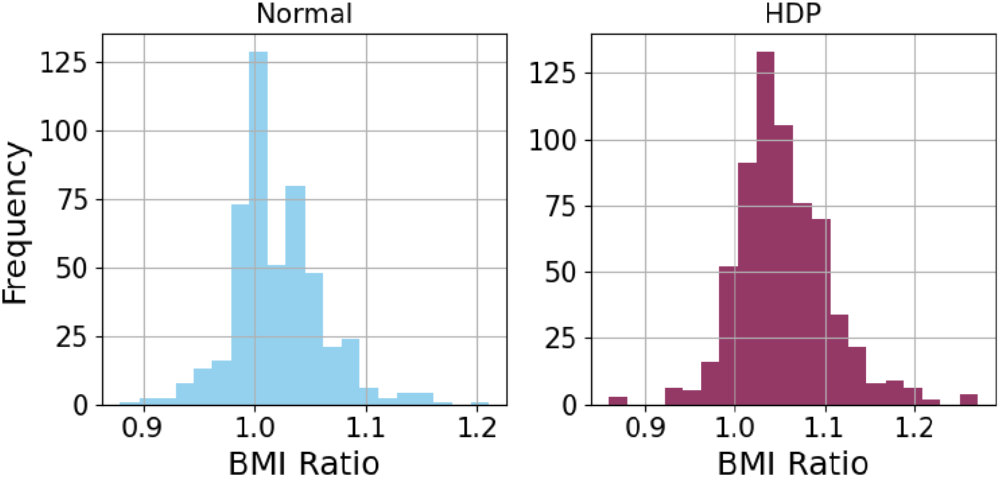
Histogram of BMI Ratio from second to first trimester based on diagnostic label.

### E. Machine Learning Classification Models

In this study, different ML methods were employed to evaluate their efficacy for the early prediction of HDPs. The models used include Extreme Gradient Boosting (XGBoost), Support Vector Machine (SVM), k-nearest Neighbors (k-NN), Multilayer Perceptron (MLP), Random Forest (RF), and voting algorithm. Each model was chosen based on its ability to handle complex, non-linear relationships and produce robust predictions in healthcare data. The models were trained using 5-fold cross-validation to provide a reliable estimate of performance. Class weights were applied to emphasize the importance of detecting HDPs, ensuring that the model gives more attention to the clinically significant class. The parameter settings for each model are detailed in Table VIII.

### F. Evaluation Metrics

The performance of the provided ML models was evaluated across various metrics to facilitate comparison with other studies. The metrics used in this study are as follows:

- **Accuracy**: The proportion of correctly predicted records.
- **Precision (Positive Predictive Value (PPV))**: The proportion of correctly predicted positive records out of all predicted positive records.
- **Recall (Sensitivity)**: The proportion of actual positive records that were correctly predicted.
- **F1-Score**: The harmonic mean of precision and recall, providing a balance between the two when dealing with imbalanced datasets.
- **Specificity**: The proportion of actual negative records that were correctly predicted.
- **Negative Predictive Value (NPV)**: The proportion of correctly predicted negative records out of all predicted negative records.
- **Receiver Operating Characteristic (ROC)**: A curve that plots the true positive rate (recall) against the false positive rate (1-specificity), allowing visualization of the model’s diagnostic ability. We reported the area under the curve as the AUROC value.
- **SHAP value analysis**: SHAP (SHapley Additive exPlanations) values are used to interpret each feature’s contribution to the model’s predictions. This analysis provides insight into how individual features influence the output, helping to explain the model’s decision-making process.

By defining these metrics, we ensure a comprehensive evaluation of the ML models, capturing both their ability to correctly predict positive and negative records. Additionally, we should specify that the reported results are based on the mean and standard deviation (SD) calculated from the 5-fold cross-validation.

## III. RESULTS

Table IX summarizes the performance of the machine learning models. The RF classifier demonstrated the best overall performance, achieving an accuracy of 0.74, an F1-score of 0.77, and an AUROC of 0.80 in classifying Normal and HDPs cases. Fig. 7 and 8 show the confusion matrix and ROC and PRC plots, respectively. The RF model’s superior performance can be attributed to its ability to handle complex, non-linear relationships and robustness against overfitting as well as its ensemble nature, which aggregates the predictions of multiple decision trees to improve generalization and reduce variance. Notably, the standard deviations for all metrics were below 0.05, underscoring the model’s robustness and consistency across different folds of cross-validation. Additionally, the results of the best model using only time-varying features are reported. The results showed minimal changes in most performance metrics. In terms of sensitivity, the voting algorithm outperformed the RF and all other classifiers, demonstrating a higher ability to correctly identify records of HDPs.

**Fig 7:**
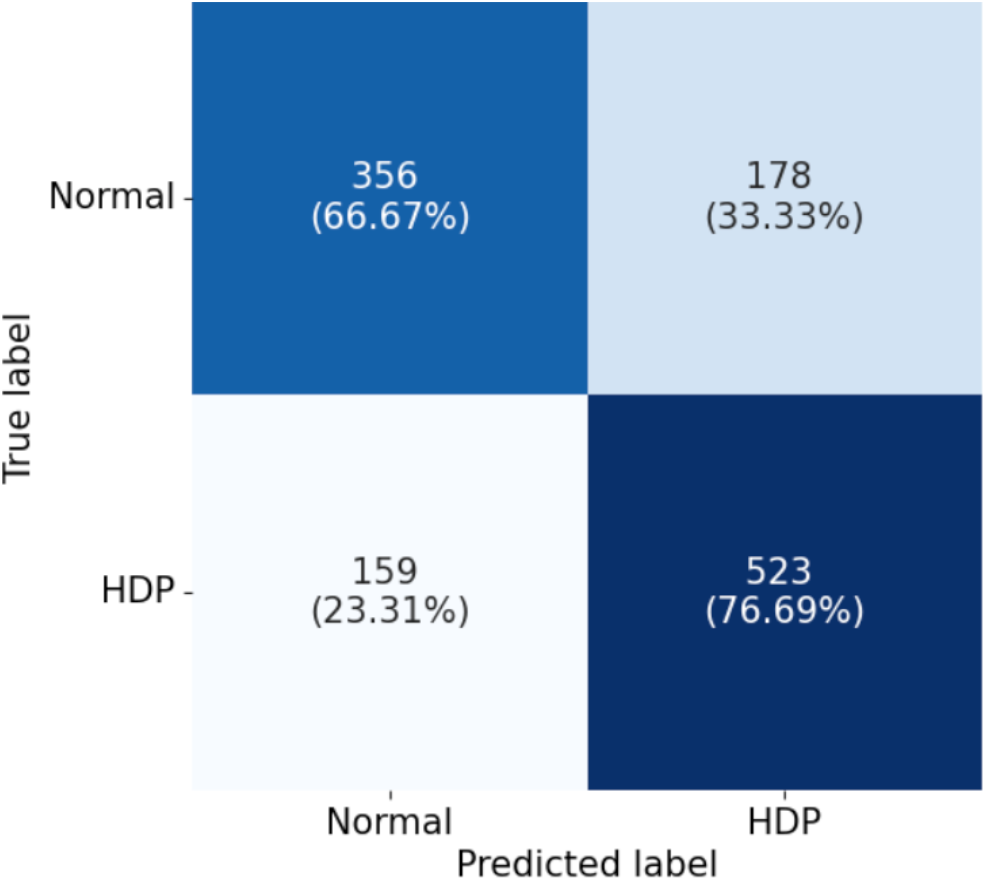
Confusion matrix of the Random Forest model for classifying normal versus Hypertensive Disorders of Pregnancy (HDPs), using features extracted from blood pressure measurements, body mass index values recorded during the first and second trimesters, and maternal demographic information (Value and percentage).

**Fig 8:**
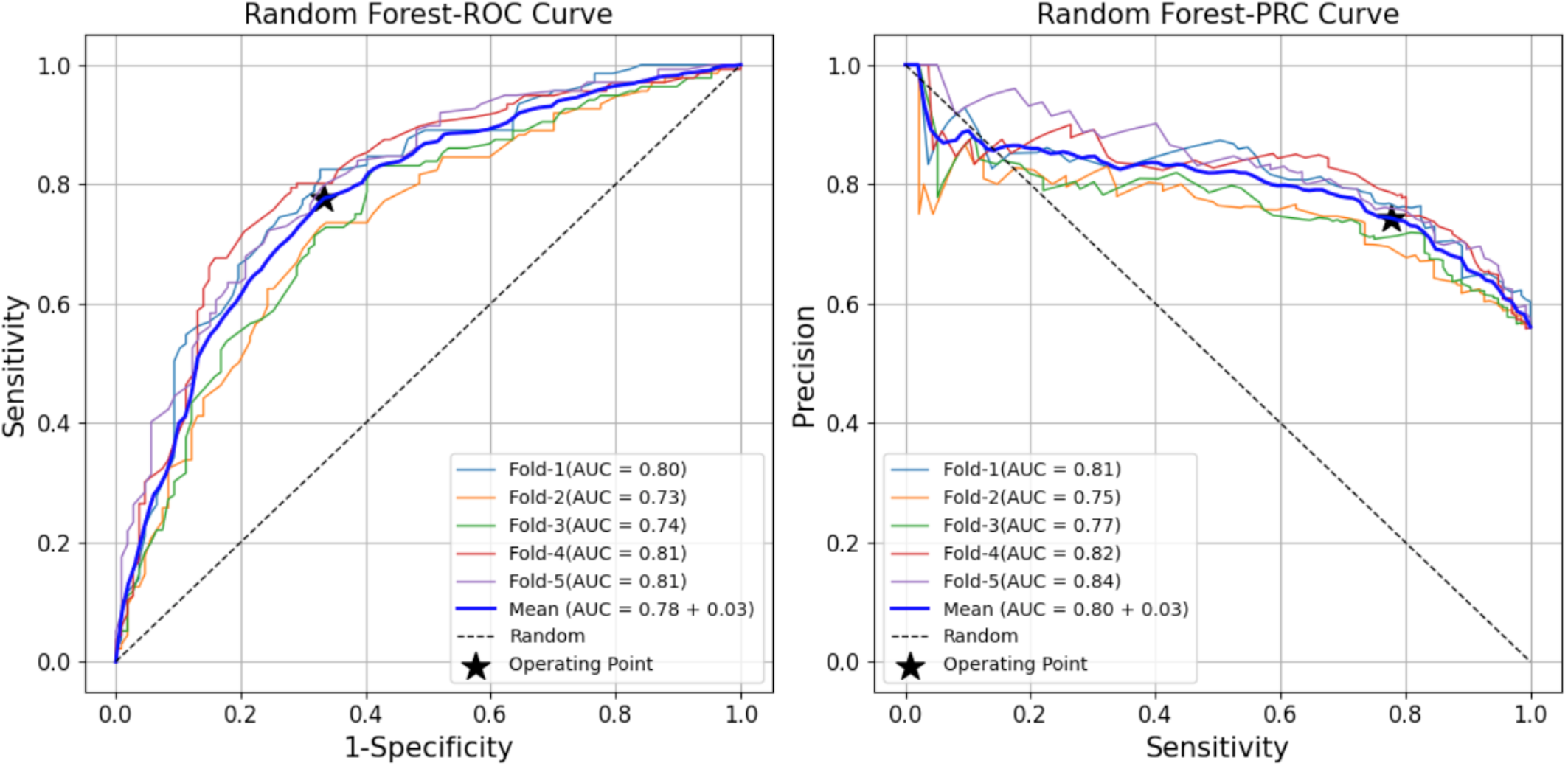
Receiver Operating Characteristic (ROC) and Precision-Recall Curve (PRC) curves of the random forest model for classifying Normal versus Hypertensive Disorders of Pregnancy (HDPs), using features extracted from blood pressure measurements, body mass index values recorded during the first and second trimesters, and maternal demographic information.

Fig. 9 presents the SHapley Additive exPlanations (SHAP) summary plot, providing a comprehensive visualization of each feature’s contribution to the model’s predictions. Based on the SHAP values, the following features have the highest impact on the model: the difference in mean BMI values between the second and first trimesters, the ratio of mean BMI between the second and first trimesters, the mean BMI during the second trimester, the mean BMI in the second trimester, the mean DBP in the first trimester, and the mean MAP in the second trimester. These features are associated with an increased likelihood of being classified as HDPs.

**Fig 9:**
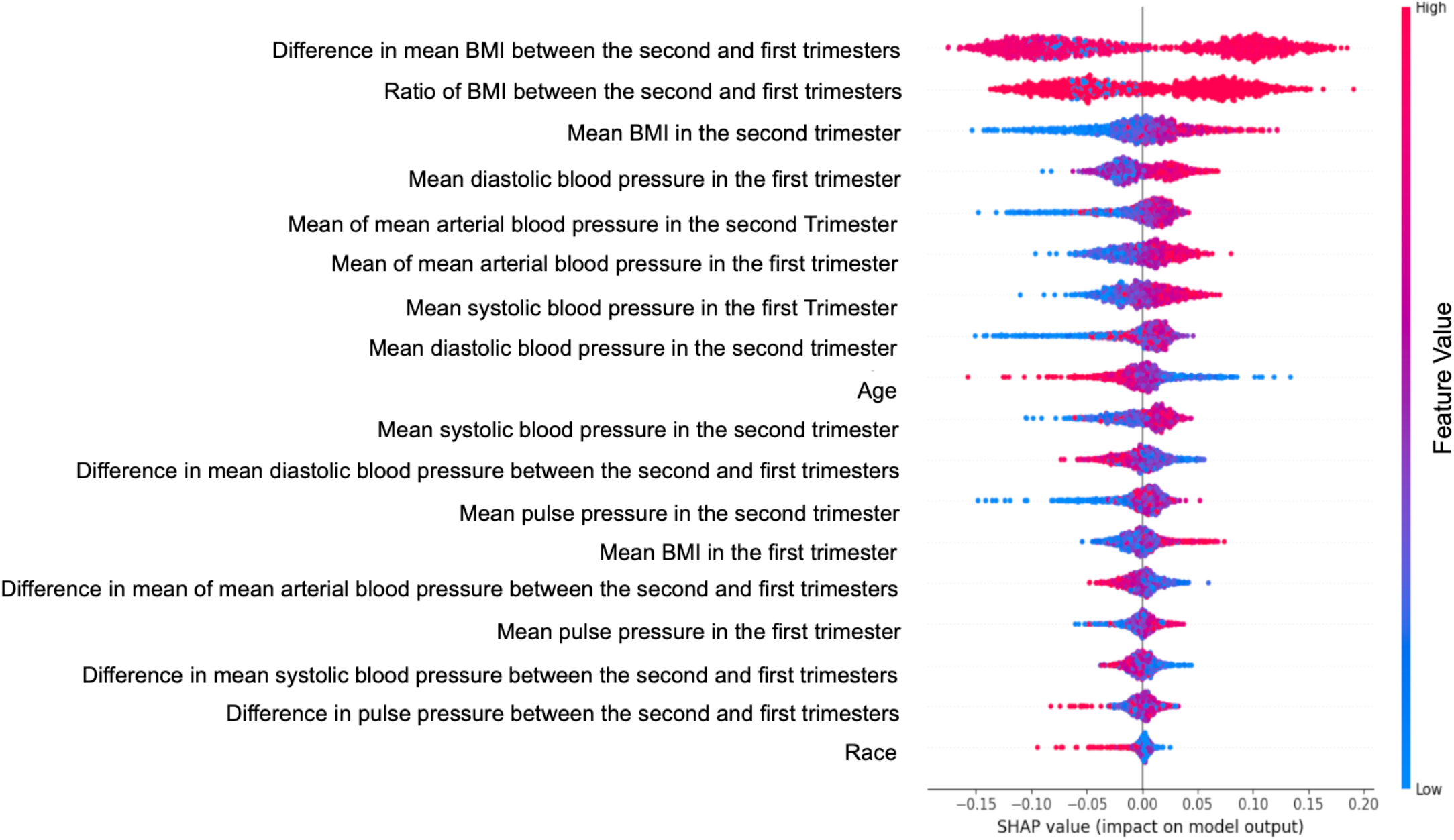
SHAP value of the random forest model for classifying Normal versus Hypertensive Disorders of Pregnancy (HDPs), using features extracted from blood pressure measurements, body mass index values recorded during the first and second trimesters, and maternal demographic information.

In the following, we compare our results with those from other studies. Table X summarizes the performance of various models based on key evaluation metrics. Compared to the model developed by Li et al. [42], which extracted features during the early second trimester, including BMI and BP values, laboratory data, demographic information, and medical history, our model demonstrates a superior F1-score and comparable precision. Although their model’s high accuracy is notable, it is important to consider that their dataset has a much lower prevalence of HDPs (4.3%) compared to the dataset in this study (56%). Additionally, our model uses only 18 features, whereas Schmidt et al. [41] developed a model with 114 features, incorporating biomarkers such as soluble fms-like tyrosine kinase-1, placental growth factor, and sonography data from both the second and third trimesters. While models trained with feature sets from the third trimester generally perform well [55], [56], they may miss opportunities for early intervention during pregnancy. Finally, we compared the results of our model with the study by Yequn Chen et al. [45], which used imbalanced datasets with only 3% HDPs records and included features from biochemical analysis, life stress levels, folic acid supplement intake, and medical history. However, our model not only achieved a close AUROC score but also demonstrated better sensitivity. In conclusion, our model, with its streamlined feature set, offers a practical and more effective approach for the early prediction of HDPs.

## IV. DISCUSSION

This study demonstrates robust performance in classifying HDPs defined based on both ICD-9 and ICD-10 codes, making the model highly relevant for clinical applications. Specifically, it can predict HDP in patients who are initially low-risk, with normal blood pressure before pregnancy, but later develop HDP. While our dataset includes records collected before the adoption of ICD-10, limiting our analysis to ICD-9 codes, this approach enhances the model’s generalizability and ensures compatibility with current clinical standards. To further enhance the model’s accuracy and applicability, it would be valuable to repeat this process with a dataset incorporating ICD-10 codes, ensuring alignment with the latest diagnostic standards.

The SHAP value analysis showed that the model considers race to be less important for predicting HDPs compared to other features. This suggests that while race may contribute to health disparities in maternal outcomes, the model prioritizes features such as BMI and BP measurements. Additionally, the dataset is relatively homogeneous in terms of race, and it includes pregnancies with visits available in both the first and second trimesters. This finding could be indicative of the model’s focus on more direct, physiologically relevant variables that are strong indicators of HDPs risk. However, the role of race in pregnancy outcomes should not be overlooked, as it may still have indirect effects on health outcomes due to factors like socioeconomic status and access to healthcare, which are not directly captured in the model.

The dataset utilized in this study was collected from Georgia, and its racial composition reflects the demographic characteristics of the region. Consequently, the dataset lacks sufficient diversity to capture the full spectrum of BP variations across different racial groups of pregnant women. This feature might affect the generalizability of our results and should be considered when evaluating the broader applicability of the model. Regarding age, more than 80 percent of maternal records fall within the age range of 25-39, and the distribution of records within these two groups shows a degree of balance. However, SHAP values indicate that with increasing age, the probability of being classified as Normal increases, while decreasing age is associated with a higher probability of HDPs. This observation is consistent with the dataset’s age distribution, as indicated by Table VI, which shows that the percentage of Normal records is about twice that of HDPs records in the age range of 40-56 years, while the percentage of HDPs records in the 16-24 age range is about 1.5 times that of Normal records. Thus, similar to race and ethnicity, it appears that we lack sufficient records in the youngest and oldest age groups. Addressing this limitation in future studies could enhance the robustness and generalizability of the model.

In addition, in this study, we categorized the data based on trimesters and calculated the mean BP and BMI values for the relevant weeks within each trimester. By increasing the frequency of measurements, such as conducting assessments weekly, the reliability of the extracted features from the dataset is expected to improve. Consequently, this approach enhances the model’s predictive accuracy, thereby providing a valuable tool for the early detection of HDPs.

## V. FUTURE WORKS

Future research should aim to validate the model with larger datasets and investigate its integration into clinical workflows to support its applicability and effectiveness further. Additionally, for future work, we propose integrating this model to the mhealth systems [57], [58] to facilitate collecting weekly or monthly BP and BMI measurements during pregnancy. Providing BP measurement devices and weight scales, which are economical methods and commonly available in most households, could facilitate early intervention, making this approach practical and accessible for broader use.

Furthermore, this approach would help mitigate sources of bias, such as masked or white-coat hypertension, by enabling more consistent and frequent monitoring in the comfort of the patient’s home.

## VI. CONCLUSION

This study developed an ML model for the early prediction of HDPs, specifically preeclampsia and GE, utilizing BP and BMI data from only the first and second trimesters. By incorporating innovative features such as the BMI ratio from the second to the first trimester, the difference in BMI values between the first and second trimesters, and the mean DBP and MAP in the first trimester, alongside the mean SBP in the second trimester, the model demonstrated a notable accuracy of 0.74. This approach not only reduces the complexity and cost of data collection but also makes the model highly practical for use in low-resource settings by relying on accessible BP and BMI measurements. The promising results suggest that this model has the potential to significantly improve early detection, intervention, and management of HDPs.

## Data Availability

All data produced in the present study are available upon reasonable request to the authors

## ACKNOWLEDGMENTS

This work was supported by a Google.org AI for the Global Goals Impact Challenge Award, the National Institutes of Health, the Fogarty International Center and the Eunice Kennedy Shriver National Institute of Child Health and Human Development, under grant # 1R21HD084114 and 1R01HD110480. G. D. Clifford is also partially supported by the National Center for Advancing Translational Sciences of the National Institutes of Health under Award # UL1TR002378. N. Katebi is funded by a PREHS-SEED award grant # K12ESO33593. The content is solely the responsibility of the authors and does not necessarily represent the official views of the National Institutes of Health.

